# Estimating the impact of Angelman syndrome on parental productivity in Australia using productivity-adjusted life years

**DOI:** 10.1101/2021.06.03.21258279

**Authors:** Sally L Hartmanis, Emma K Baker, David E Godler, Danny Liew

**Affiliations:** School of Public Health and Preventive Medicine, Monash University, 553 St Kilda Road, Melbourne, Victoria 3004, Australia; Diagnosis and Development, Murdoch Children’s Research Institute, Royal Children’s Hospital, 50 Flemington Road, Parkville, Victoria 3052, Australia; Faculty of Medicine, Dentistry and Health Sciences, Department of Paediatrics, University of Melbourne, Parkville, Victoria 3010, Australia; School of Psychology and Public Health, La Trobe University, Plenty Road and Kingsbury Drive, Bundoora, Victoria 3086, Australia

**Author notes:** **Corresponding author** Sally L Hartmanis, School of Public Health and Preventive Medicine, Monash University, 553 St Kilda Road, Melbourne, Victoria 3004 Australia.

**Keywords:** Angelman syndrome, Intellectual disability, Parents, Productivity burden, Cost-of-illness, Economic impact

## Abstract

**Background:** Angelman syndrome (AS) is a rare genetic condition characterised by global developmental delay, including severe to profound intellectual disability. The parents of persons with AS experience increased stress, anxiety and depression. This impacts parents’ career choices and productivity.

**Aims:** To estimate, for the first time, the total productivity lost by the parents of persons with AS over a 10-year period in Australia and the corresponding cost to society.

**Methods and procedures:** A cost-of-illness model with simulated follow-up over a 10-year period was developed, with 2019 as the baseline year, facilitated by a Markov chain of life tables. The prevalence of persons with AS and their parents, the productivity-adjusted life years (PALYs) lost by parents, and the cost to society were estimated. Key data were obtained from a prospective cohort of AS families, peer-reviewed literature, and publicly available sources.

**Outcomes and results:** The base-case productivity burden borne by the estimated 330 living parents of the 428 prevalent-persons with AS totalled AUD$45.30 million, corresponding to a loss of 38.42% of PALYs per-parent.

**Conclusions and implications:** Caring for a child with AS has a significant impact on the productivity of affected parents, with a large associated impact on the broader Australian economy.

**What this paper adds?:** Persons with AS require lifelong care and support. Consequently, AS results in a significant socioeconomic impact, borne both by the healthcare system and affected families. This is the first known study to estimate the total impact of caring for a child with AS on parental productivity, as well as the first study known to estimate the PALYs lost by a parental or caregiver population. This study found that caring for a child with AS has a significant impact on the productivity of affected parents, with a large associated impact on the broader Australian economy. At present, the supports available to persons with AS and their families include sleep aids and behavioural therapy. In future, specific therapeutic treatments for AS may become available, with trials underway at present investigating the efficacy and effectiveness of gene therapies for AS. As such, evidence regarding the total socioeconomic impact, including the parental productivity burden, attributable to AS is needed to inform future funding decisions.

## Introduction

Angelman syndrome (AS) is a rare genetic condition characterised by global developmental delay, including severe to profound intellectual disability, absent speech and motor function impairment (Khan et al., 2019b, 2019a; Wheeler et al., 2017). A “complex and variable” behavioural profile is also observed including sleep disturbances, hyperactivity, excessive smiling and laughing, and repetitive and aggressive behaviours (Khan et al., 2019b, 2019a; Wheeler et al., 2017). Persons with AS can experience multiple comorbidities, including epilepsy, cerebral palsy, scoliosis, and blindness and other visual impairments, as well as autism features (Baker et al., 2018; Jørgensen et al., 2019a; Wheeler et al., 2017).

Estimates of AS prevalence vary greatly from 1 in 8,300 to 1 in 86,250 (Jørgensen et al., 2019b). Furthermore, many of the studies reporting population prevalence have estimated this using small samples of persons with intellectual disability, followed by extrapolation to the total population (Wheeler et al., 2017). The epidemiological characteristics of AS do not appear to differ by race (Luk & Lo, 2016). This large degree of variability may be attributed to several factors. Specifically, the absence of a newborn screening program for AS to allow for estimates on cohorts of unbiased ascertainment, overlapping phenotypic characteristics with other syndromes, issues pertaining to the accuracy of diagnostic methods, the lack of genetic confirmation for approximately 10% of cases, and historically low levels of awareness among clinicians (Õiglane-Shlik et al., 2006; Thomson et al., 2006; Tones et al., 2018; Wheeler et al., 2017). Many of these factors mean that children with AS, and their families, are often subject to a ‘lengthy diagnostic odyssey’ with most only receiving a diagnosis between the ages of one and five years (Godler et al., 2022; Wheeler et al., 2017).

Persons with AS require lifelong care and support, with the average lifespan of a person with AS being reduced by approximately 10 to 15 years (Coppus, 2013; Dagli et al., 2011; Khan et al., 2019a). Consequently, AS results in a significant socioeconomic impact, borne both by the healthcare system and affected families (Wheeler et al., 2017). Specifically, persons with AS require a high-level of specialist medical care within both an outpatient and inpatient setting, with the most frequent causes of hospitalisation comprising care for oral-dental issues, seizures, orthopaedic problems, and acute respiratory disorders (Domínguez-Berjón et al., 2018; Khan et al., 2019b). This is most pronounced among young children with AS, who tend to require a greater number of surgeries and hospitalisations, and who need to spend more time in hospital once admitted (Khan et al., 2019b). Furthermore, the care required by persons with AS when undergoing routine health-related procedures often involves a significantly greater use of resources due to the need for additional interventions (Thomson et al., 2006). These can include, for example, a general anaesthetic when visiting the dentist for a routine teeth cleaning service (Thomson et al., 2006). In addition, persons with AS often require many pharmaceuticals either for directly treating the symptoms of their condition or one of the recognised comorbidities of AS, with almost 80% of children aged six years requiring at least one medication. This is despite the fact that there is no specific therapeutic treatment for AS (Jørgensen et al., 2019a; Khan et al., 2019b, 2019a; Wheeler et al., 2017). Furthermore, the parents of persons with AS are at significant risk of stress, fatigue, and mental health problems. This risk is greater than that experienced by the parents of persons with autism, Cornelia de Lange syndrome and Cri du Chat syndrome (Griffith et al., 2011). This impact is particularly notable for mothers of children with AS (Griffith et al., 2011; van den Borne et al., 1999; Wulffaert et al., 2010). However, the potential positive impacts arising from caring for a child with AS are less well understood (Wheeler et al., 2017).

Despite the above considerations, there remains limited evidence regarding the productivity impact borne by the parents of persons with AS, with limited literature on the adverse effects on parents’ employment and productivity (Grieco et al., 2019; Khan et al., 2019b; Thomson et al., 2017; Wheeler et al., 2017; Willgoss et al., 2020). This impact can be quantified using productivity-adjusted life years (PALYs), which is a measure of the productivity burden imposed by a given condition (Ademi et al., 2021). PALYs are useful for estimating and communicating productivity impacts because they can be compared across conditions and populations to inform policy and funding decisions (Ademi et al., 2021). PALYs are a function of the life years lived by a given population and their corresponding productivity index. The productivity index ranges from zero (completely unproductive) to one (completely productive) and is a measure of workforce participation, time off work (absenteeism), and reduced productive output while at work (presenteeism) (Ademi et al., 2021; Giovannetti et al., 2009). Each of these components are known to impact the parents of persons with AS (Grieco et al., 2019; Khan et al., 2019b; Thomson et al., 2017; Wheeler et al., 2017; Willgoss et al., 2020).

The present study aimed to estimate, for the first time, the total productivity lost by the parents of persons with AS over a 10-year period in Australia and the corresponding cost to society. It was hypothesised that caring for a person with AS has a significant impact on the productivity of affected parents, with a large associated impact on the broader Australian economy.

## Methods

The study was approved by the Royal Children’s Hospital Human Research Ethics Committee (reference number 33066) and the Monash University Human Research Ethics Committee (reference number 2021-25930-55113). All parents/caregivers provided written informed consent and participants who were deemed cognitively able also provided written informed consent.

### 1.1. Modelling approach

The productivity burden borne by the parents of persons with AS in Australia was estimated using cost-of-illness modelling with simulated follow-up over a 10-year period with 2019 as the baseline year, facilitated by a Markov chain of life tables. This involved estimating the prevalence of persons with AS and their parents, the PALYs lost by parents, and the associated cost to society. The model structure is depicted in Figure 1.

**Figure 1.**
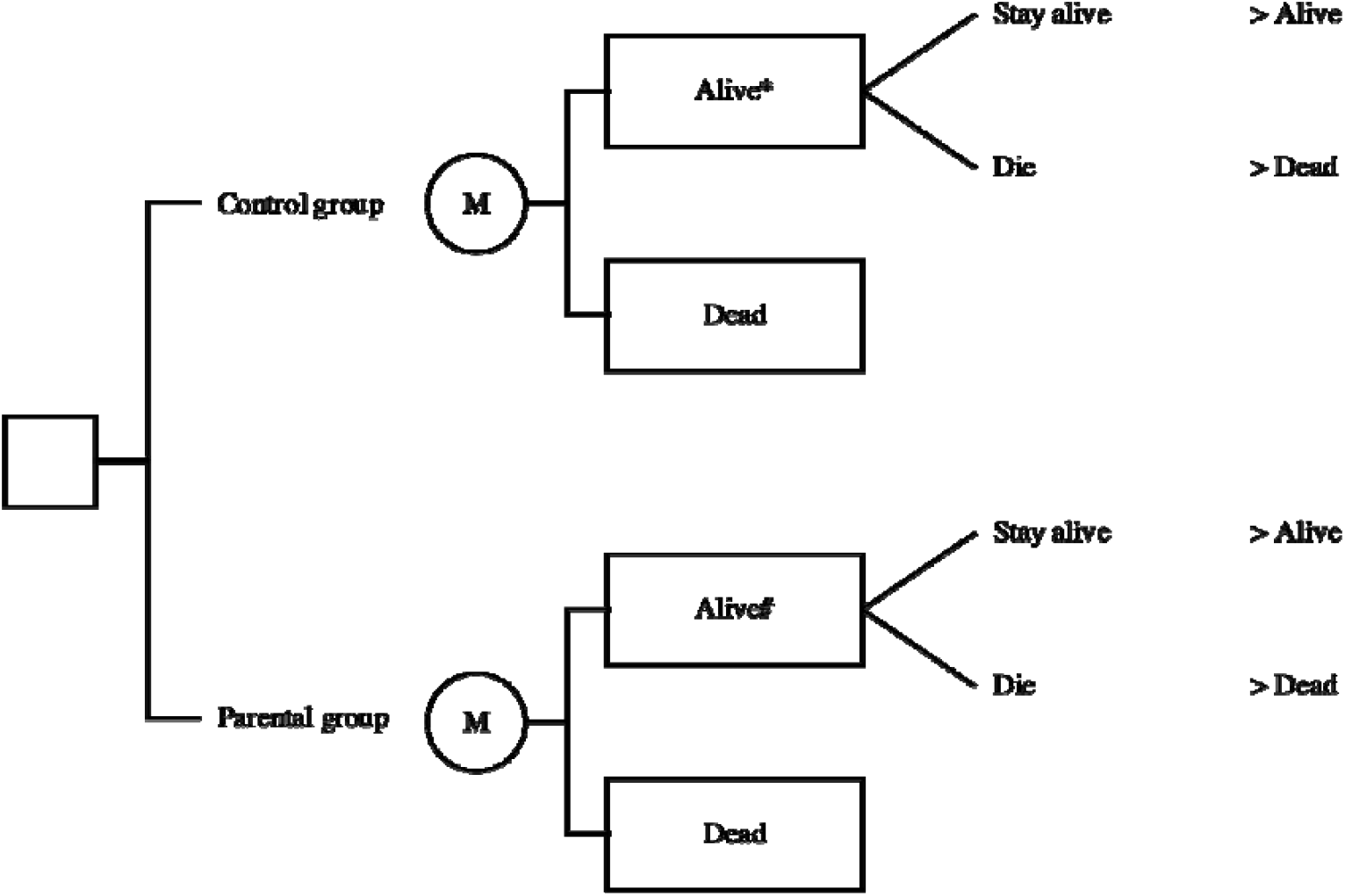
Structure of the cost-of-illness model developed to estimate the productivity burden borne by the parents of persons over a 10-year period in Australia. *Productivity of the aged and sex matched, general Australian population #Productivity altered due to caring for persons with AS

Key data were obtained from a prospective cohort of families of children with AS who participated in a natural history study of chromosome 15 imprinting disorders (Baker et al., 2018, 2021), peer-reviewed literature, and publicly-available data reported by the Australian Bureau of Statistics (ABS). The data analyses and supporting assumptions are discussed in the following sections, with further detail provided in Tables A.1 and A.2.

Approaches were aligned to the applicable sections of the Consolidated Health Economic Evaluation Reporting Standards (CHEERS) statement for health interventions (Husereau et al., 2013).

### 1.2. Data analysis

#### 1.2.1. Prevalence of AS

The prevalence of AS in Australia was estimated by determining the population prevalence of AS, the age and sex distribution of prevalent AS, and the average life expectancy of persons with AS.

Few studies reporting the population prevalence of AS in Australia were identified and variation across the reported estimates in other countries was significant (Al Salloum et al., 2015; Godler et al., 2022; Jørgensen et al., 2019a; Luk & Lo, 2016; Mertz et al., 2013; Õiglane-Shlik et al., 2006; Petersen et al., 1995; Thomson et al., 2006). As such, results were estimated for a lower, base-case, and upper prevalence scenario to account for the underlying uncertainty in the prevalence of AS within Australia. These estimates were applied to the total number of Australians in 2019 reported by the ABS to derive the total number of prevalent AS cases (*National, State and Territory Population*, 2020). The age and sex distribution of prevalent AS was assumed to follow the same distribution as that of prevalent disability in Australia in 2018 as reported by the ABS (*Disability, Ageing and Carers*, 2018). The average life expectancy of persons with AS was assumed to be 70 years (Coppus, 2013; Dagli et al., 2011).

The base-case estimate of prevalence was obtained from an Australian retrospective record review which identified 26 persons with AS having been born in Western Australia during the 50 years that Disability Services Commission (DSC) records were kept, corresponding to a live birth incidence of 1:40,000 (0.0012%) among a birth population of 1.05 million (Thomson et al., 2006). This estimate was selected as the base-case estimate due to the proximal relationship of incidence and prevalence in rare disease, and the crossover of the Thomson et al (2006) study population (Western Australians) with the target population of this study (Australians). The lower prevalence estimate was obtained from a retrospective registry review in Denmark, which identified 80 patients with AS out of the 6.90 million patients in the Danish National Patient Registry (DNPR) from 1994 to 2015, corresponding to a prevalence of 1:86,250 (0.001%) (Jørgensen et al., 2019a). The upper prevalence estimate was obtained from a study evaluating the feasibility of screening for chromosome 15 imprinting disorders in infants born in Victoria, Australia which reported a prevalence of 1:8,290 (0.012%) for infants born in 2011 (Godler et al., 2022).

#### 1.2.2. Prevalence of the parents of persons with AS

The prevalence of the parents of persons affected by AS was estimated by determining the average parental age during the year that each child with AS was born. This was estimated by deriving a weighted average of maternal and paternal age, respectively, for the required years using data reported by the ABS (*Births*, 2019). This approach assumed that the parents of persons with AS had the same average age as all Australians when their child was born. In addition, this approach assumed that each person with AS had one biological mother and one biological father who experienced productivity impacts attributable to AS.

#### 1.2.3. PALYs lost

The PALYs lost by this prevalent group of parents were estimated by comparing the PALYs lived by this group with the PALYs lived by a simulated control group. This simulated control group allowed for the estimation of a counterfactual scenario in which the parents of persons with AS did not have a child with AS and, therefore did not experience any productivity impacts attributable to AS. It was assumed that the mortality rate of persons with AS had a negligible impact on the AS attributable productivity impacts experienced by parents. PALYs accrued beyond the first year of the modelled simulation were discounted at 5.0% per annum (Larg & Moss, 2011).

The number of life years lived by the parental and simulated control groups were estimated via the construction of a Markov chain of age and sex specific life tables to simulate the progress of the parental and control groups over a 10-year period, adjusted for background mortality (*Deaths*, 2019). Each group was followed until a maximum age of 70 years. Separate life tables were constructed for 20 age and sex sub-cohorts, with age stratified into 10 five-year age groups from 20-24 years to 65-69 years. The starting age in each sub-cohort was the mid-point of that age group. The 20-69 years age range was selected to reflect the ages during which people were commonly engaged in paid employment. It was assumed that there was no mortality attributable to being the parent of a person with AS, meaning life years lived in the parental and simulated control groups were the same. As such, the difference in the PALYs lived by each group was driven by their respective productivity indices.

The productivity index of the parental group was derived by estimating this group’s average workforce participation, and the level of absenteeism and presenteeism attributable to having a child with AS. Uncertainty in these inputs was accounted for in sensitivity analyses in which the estimated parental average workforce participation, and attributable absenteeism and presenteeism were varied by +/-20%.

The average workforce participation of the parents of persons with AS was estimated using data collected from a prospective cohort of families of children with AS who participated in a natural history study of chromosome 15 imprinting disorders (Baker et al., 2021). A Client Service Receipt Inventory (CSRI) modified to be suitable for AS assessments in Australia and Participant Developmental and Medical Anamnesis (PDMA) questionnaire were distributed to 28 parents of persons with AS (Baker et al., 2021). The parents responded to the questionnaires on behalf of themselves and their spouse/partner. A sub-set of these data were used to derive the average workforce participation of 50 individual parents of persons with AS. As such, it was estimated that average workforce participation was 97.3% for fathers and 47.4% for mothers. This approach assumed that the workforce participation reported by this sample size was reflective of the average workforce participation of the parents of persons with AS across all age groups included in the analysis.

No studies estimating absenteeism and presenteeism attributable to being the parent of a person with AS were identified. Therefore these inputs were estimated by proxy using the results from a study of the caregiver burden attributable to Dravet syndrome (DS); a rare genetic disorder characterised by epilepsy and developmental delay, in which caregiver productivity impairment was measured using a subset of the Work Productivity and Activity Impairment (WPAI) questionnaire; a tool which has been validated for eliciting attributable caregiver productivity loss (Ademi et al., 2021; Campbell et al., 2018; Giovannetti et al., 2009). Of parents remaining in the workforce, their mean time missed from work each week ranged from 6.9 to 7.4 hours, while their productive output at work was impacted by 39.1 to 76.9% (Campbell et al., 2018). The mid-points of each of these ranges (7.2 hours and 58.0%, respectively) were used as inputs in the analysis.

The productivity index of the simulated control group was derived by estimating the average workforce participation of the general population. This was estimated using age and sex specific ABS employment data for 2019 (*Characteristics of Employment*, 2020). This dataset includes data pertaining to adults with and without children, including children with disabilities. As such, it was assumed that the data approximated the average workforce participation of the control population. Absenteeism and presenteeism were not estimated for the general population because *relative* absenteeism and presenteeism measures attributable to AS were applied.

#### 1.2.4. Cost to society

The cost to society resulting from this lost productive output was estimated by multiplying the number of PALYs lost within each age and sex sub-cohort included in the analysis by the average gross domestic product (GDP) per full-time equivalent (FTE) worker specific to that sub-cohort (Ademi et al., 2021; *National Income, Expenditure and Product*, 2020). GDP per FTE is a function of GDP per hour worked per person adjusted for the proportion of FTEs within each age and sex sub-cohort (*Characteristics of Employment*, 2020; *National Income, Expenditure and Product*, 2020). This assumed that the hourly contributions to GDP made by parents who worked were the same as the average for all working Australians.

## Results

### 1.3. Prevalence of persons with AS and their parents

The number of people with AS in Australia in 2019 was estimated to total 428 (lower: 198, upper: 2,069) persons (Figure 2). The number of parents of persons affected by AS in Australia in 2019 between the ages of 20 and 69 years was estimated to total 330 (lower: 153, upper: 1,594) (Figure 3).

**Figure 2.**
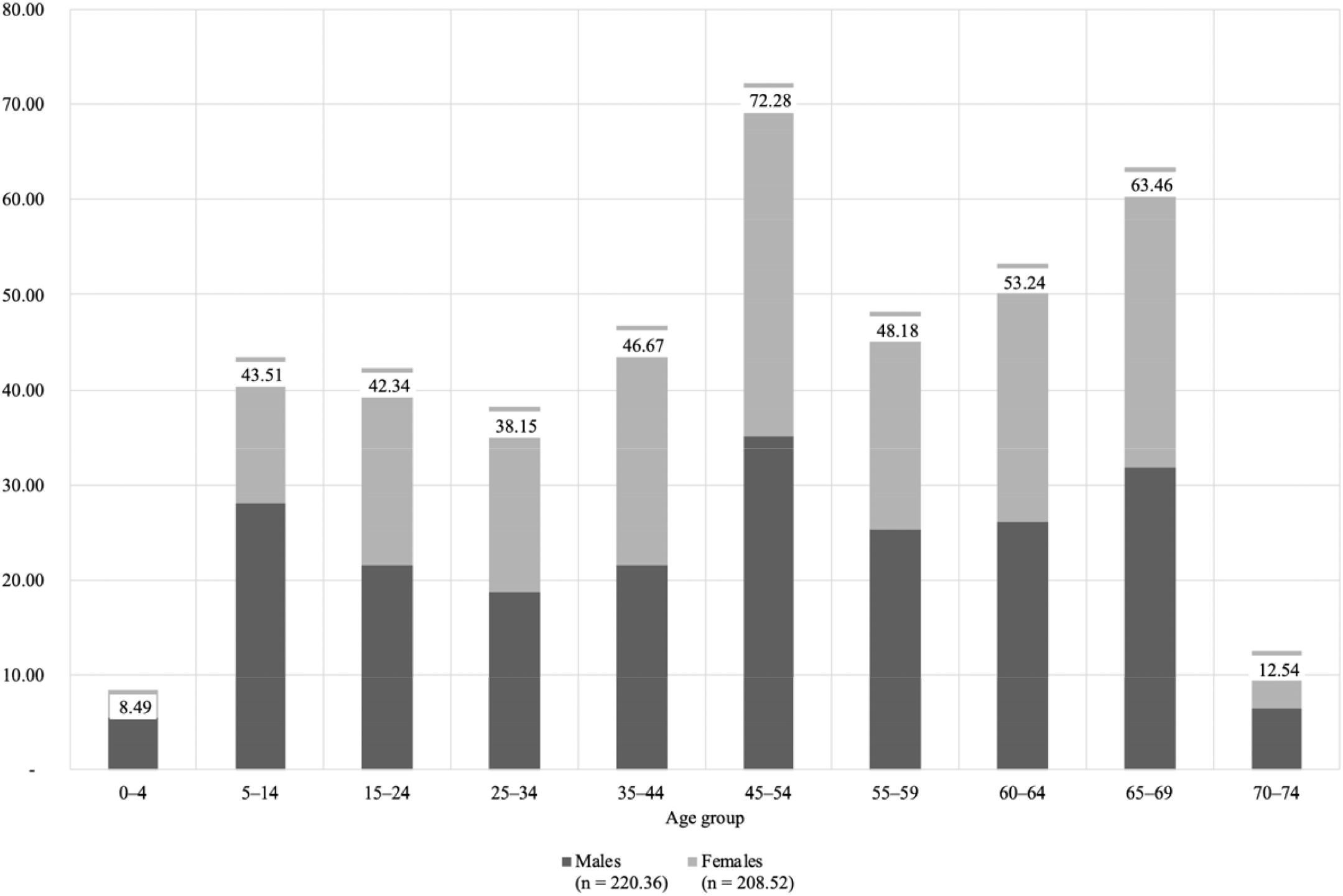
Distribution of the number of people with AS in Australia by age and sex (base-case scenario)

**Figure 3.**
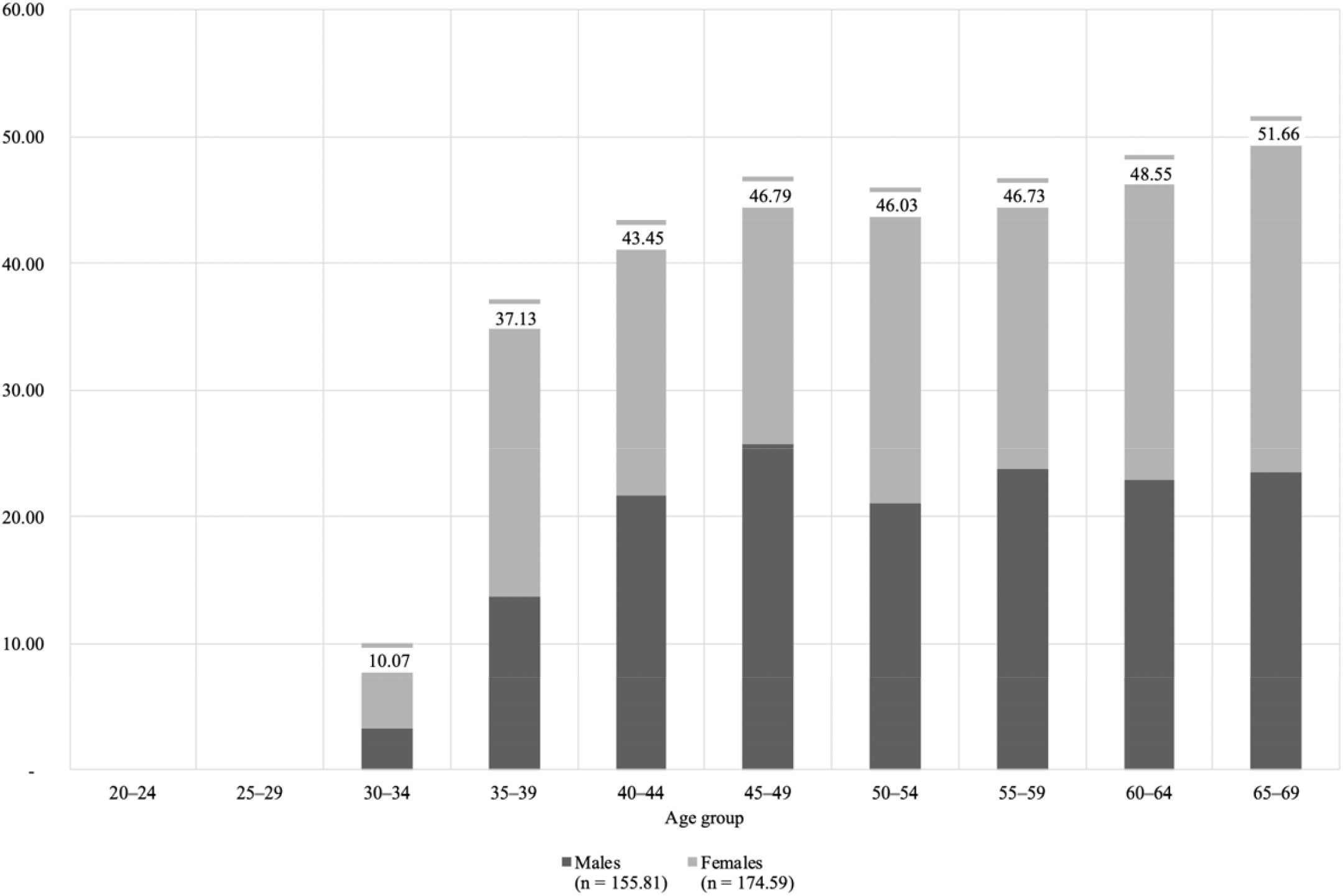
Distribution of the number of parents of people with AS in Australia by age and sex (base-case scenario)

### 1.4. PALYs lost and cost to society

The productivity burden borne by the parents of persons with AS in Australia over a 10-year period was estimated to total 183.40 (lower: 85.06, upper: 884.92) PALYs (fathers), 311.66 (lower: 144.54, upper: 1,503.76) PALYs (mothers), and 495.06 (lower: 229.59, upper: 2,388.69) PALYs (total) (Table 1). These values were discounted, as described in the Methods section. This corresponded to a loss of 25.19% of PALYs (fathers), 53.05% of PALYs (mothers) and 38.42% of PALYs (average) per-parent (Table 1). As expected, the proportion of PALYs lost was greatest between the ages of 30 and 54 years (Figure 4).

**Table 1.** The productivity burden borne by the parents of persons with AS in Australia over a 10-year period.

|  | Fathers | Mothers | Total |
| --- | --- | --- | --- |
| **PALYs lost** |  |  |  |
| Total (PALYs) | base-case: 183.40<br>(lower: 85.06, upper: 884.92) | base-case: 311.66<br>(lower: 144.54, upper: 1,503.76) | base-case: 495.06<br>(lower: 229.59, upper: 2,388.69) |
| Per parent (PALYs) | 1.18 | 1.79 | 1.50 |
| Proportion per parent (%) | 25.19 | 53.05 | 38.42 |
| **Cost to society** |  |  |  |
| Total (AUD\$, million) | base-case: 19.98<br>(lower: \$9.27, upper: \$96.42) | base-case: 25.32<br>(lower: \$11.74, upper: \$122.16) | base-case: 45.30<br>(lower: \$21.01, upper: \$218.57) |
| Per parent (AUD\$) | 128,252.16 | 145,007.16 | 137,105.94 |

**Figure 4.**
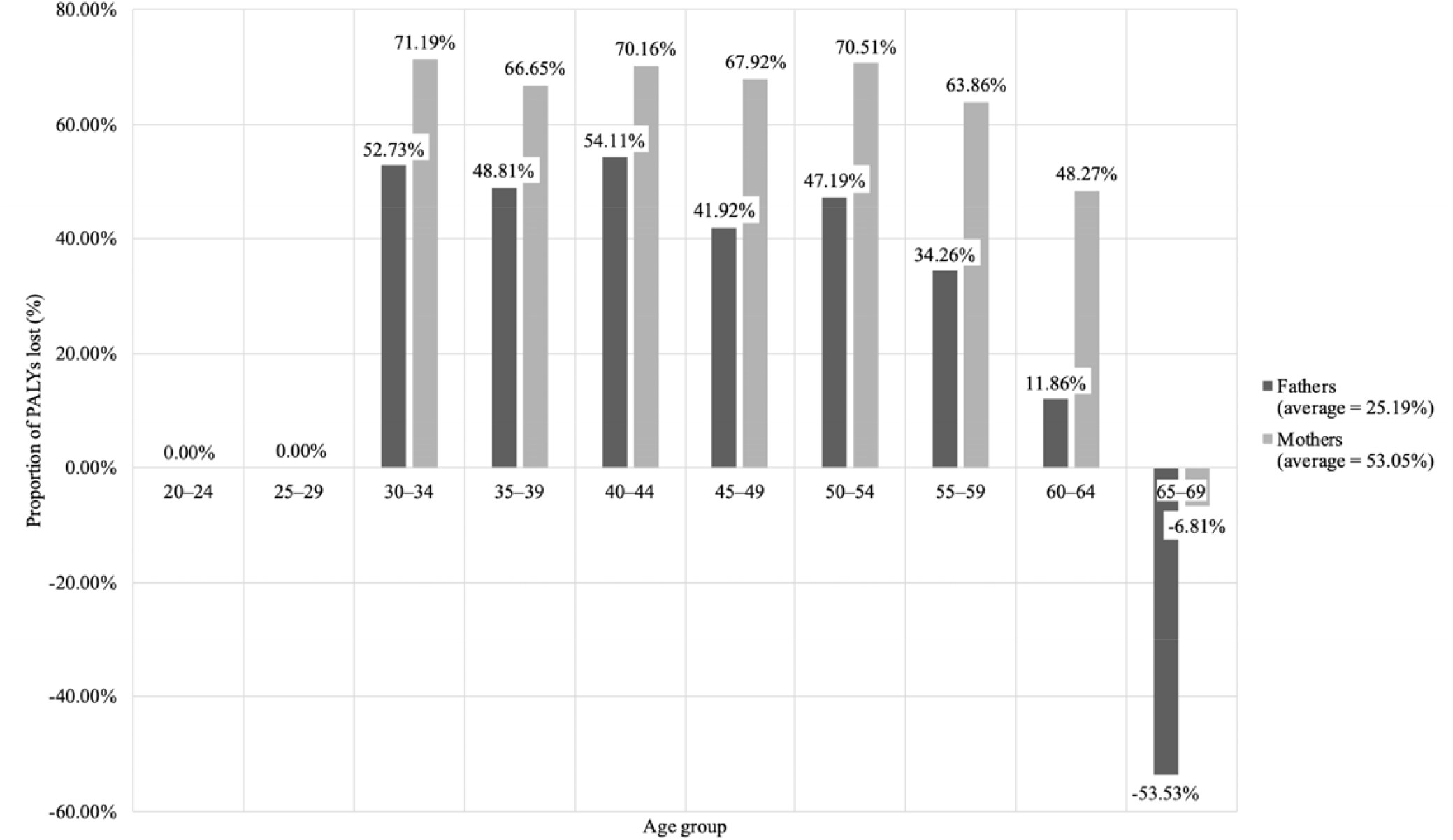
The proportion of PALYs lost by the parents of persons with AS in Australia over a 10-year period by age and sex. Note: A negative productivity loss was estimated for the 65-to-69-year age group due to the lack of age specific parental productivity indices, resulting in the parental population accruing greater PALYs in this age group than the control population.

This productivity impact corresponded to a societal cost of AUD$19.98 (lower: $9.27, upper: $96.42) million (fathers), AUD$25.32 (lower: $11.74, upper: $122.16) million (mothers), and AUD$45.30 (lower: $21.01, upper: $218.57) million (total) (Table 1). This equated to a per-parent cost of AUD$128,252.16 (fathers), AUD$145,007.16 (mothers), and AUD$137,105.94 (average) (Table 1). These values were discounted by 5%.

### 1.5. Sensitivity analyses

When reviewing the results of the sensitivity analyses across the prevalence scenarios estimated, it was evident that the paternal results are most sensitive to fluctuations in the key parental productivity index inputs (Figure 5). This is likely due to the extremely high average paternal workforce participation estimated. In addition, variations in the presenteeism inputs resulted in significant variation in the results.

**Figure 5.**
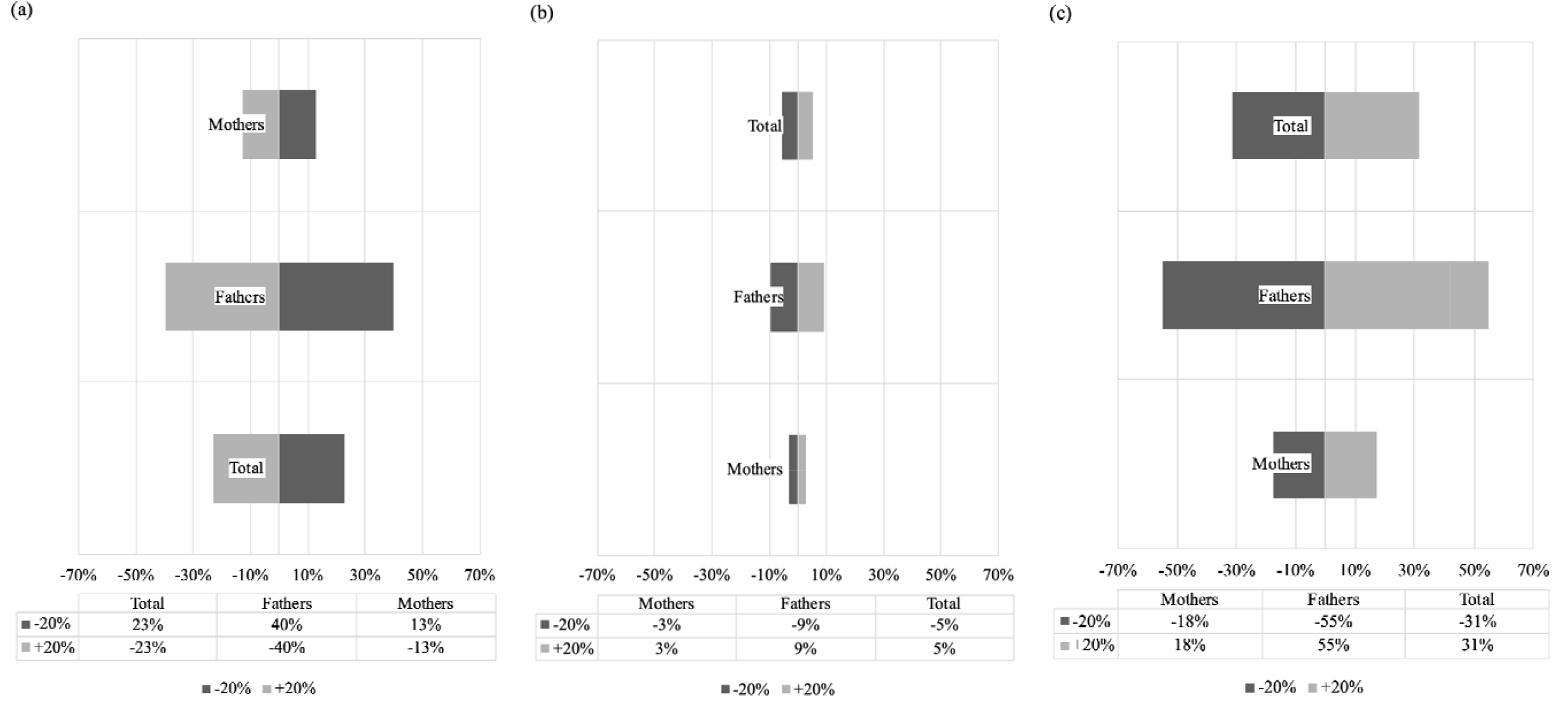
Sensitivity analyses (a) effect of +/- 20% average parental workforce participation on PALYs lost (b) effect of +/- 20% attributable absenteeism on PALYs lost (c) effect of +/- 20% attributable presenteeism on PALYs lost.

## Discussion

The present study highlights the significant impact that caring for a child with AS imposes on parents’ productivity, and the broader economy. Mothers were found to bear the bulk of this burden, while by contrast, fathers were found to have higher workforce participation than the average population. The latter may be reflective of a productivity impact in reverse, in which fathers remain in fulltime employment at greater levels than they otherwise would have. This may be due to the high costs of raising a child with AS which are not always covered by government subsidies, and may be associated with mental health and quality of life impacts not captured in the present study (Baker et al., 2021). As expected, the socioeconomic cost of this lost productivity was greatest during parents’ prime working years.

The present study fills an important gap in the literature by quantifying and monetising the productivity burden borne by the parents of persons with AS. Previous studies have qualitatively explored this topic, but did not quantify or monetise the parental productivity impact of AS (Grieco et al., 2019; Miodrag & Peters, 2015; Thomson et al., 2017; Wheeler et al., 2017; Willgoss et al., 2020). These studies found that the parents of persons with AS experience increased stress which, in turn, is associated with poor parental health. In particular, the sleep issues experienced by persons with AS, such as insomnia; wake time after sleep onset; fragmented sleep; and variability in total sleep time, are a major cause of parental stress and are also linked with higher rates of parental insomnia and daytime drowsiness (Goldman et al., 2012; Grieco et al., 2019; Wheeler et al., 2017).

Beyond stress and fatigue, the parents of persons with AS also experience increased worry, depression, anxiety, fear, frustration, irritability, loss of control, social isolation and feelings of being unsupported (Wheeler et al., 2017). This may impact parents’ relationships with their spouse/partner, other children, extended family and social network (Grieco et al., 2019; Wheeler et al., 2017). This may also be further exacerbated by the extent to which the lives of parents and the broader family unit revolve around the needs of the person with AS (Grieco et al., 2019). However, the parental impacts of caring for a person with AS are not limited to psychosocial aspects. They can extend into the physical health domain, with many parents and caregivers having experienced back and other chronic pain from lifting and/or assisting their child, as well as other injuries resulting from aggressive behaviours such as hitting, scratching and biting (Grieco et al., 2019; Wheeler et al., 2017). Furthermore, the impacts of caring for a person with AS extend into parents’ career choices, with reports of some parents having to change jobs, change careers, decrease their work hours or leave the workforce entirely (Grieco et al., 2019; Wheeler et al., 2017; Willgoss et al., 2020). In addition, those who remain in the workforce may experience greater time off work and reduced productivity while at work, as was identified in Grieco et al (2019) and estimated in this study.

Previous studies estimating the productivity impact of intellectual disability in Australia provide useful context and points of comparison (Arora et al., 2020; Baker et al., 2021; Doran et al., 2012). However, these studies each addressed slightly different research questions, and hence their results are not directly comparable to those described in the present study. Baker et al (2021) estimated the productivity losses arising from the under- and un-employment of individuals with AS from an individual and government perspective. In doing so, Baker et al (2021) found that the mean income tax revenue lost due to the reduced employment of individuals with AS totalled AUD$19,367 (95% CI: $7,785-$30,949) per annum. By contrast, Arora et al (2020) sought to quantify the costs associated with intellectual disability in childhood in Australia, including costs associated with healthcare, informal care, and productivity losses. Arora et al (2020) estimated that the total cost of intellectual disability in Australia was AUD$72,027 per child and $12.5 billion per year, including $142.4 and $239.3 per month due to paid and unpaid absences from work, respectively. Arora et al (2020) also found that only 47% of parents reported being employed, but this impact was not included in the productivity losses estimated. Similarly, Doran et al (2012) estimated the cost of intellectual disability in Australia, arriving at a value of AUD$14.7 billion per year, with the opportunity cost of lost time accounting for 85% of this cost. Doran et al (2012) also found that over 70% of parents sacrifice work opportunities to accommodate their caregiving responsibilities. However, the productivity losses associated with this burden were not estimated (Doran et al., 2012). By contrast, Doble et al (2020) sought to estimate the impact of genomic testing on the total socioeconomic cost of monogenic disorders resulting in intellectual disability, including caregiver productivity costs. Doble et al (2020) found that the costs associated with intellectual disability average USD$172,000 per person per year. However, the productivity costs estimated were reported as a combined result for individuals with intellectual disability and their caregivers. As such, the caregiver productivity costs estimated by Doble et al (2020) were unable to be compared to those estimated in the present study.

These studies indicate that in addition to parental productivity costs, intellectual disability results in many other costs to society, families, and individuals. These include costs associated with healthcare, aids and modifications, community services, specialist education and day placement services, individual productivity losses (i.e., borne by the disabled person as distinct from those borne by their parents), residential and respite care, formal and informal care, and losses of wellbeing borne by the individual as well as their parents and siblings. As such, the lost parental productivity estimated in the present study is likely to be only a fraction of the total socioeconomic cost attributable to AS.

Cost-of-illness studies are used to estimate the socioeconomic impact of a disease or disability on a population (Larg & Moss, 2011). These studies can be used to raise awareness of the condition, drive investment in therapeutic research and development, and provide critical inputs to government regulatory and reimbursement decision making (Larg & Moss, 2011). As such, the present targeted cost-of-illness study could be used to inform government decisions regarding the supports that should be provided to persons with AS and their families (Khan et al., 2019b). At present, the supports available to persons with AS and their families include sleep aids and behavioural therapy (Wheeler et al., 2017). In future, supports may also include specific therapeutic treatments for AS, with trials underway at present investigating the efficacy and effectiveness of gene therapies for AS (Godler et al., 2022). Therefore, evidence regarding the total socioeconomic impact, including the parental productivity burden, attributable to AS is needed to inform future funding decisions.

Several limitations to the present study warrant mention. First, there was uncertainty regarding AS population prevalence and the parental productivity index inputs. Specifically, no studies estimating absenteeism and presenteeism attributable to being the parent of a person with AS were identified. As such, these inputs were estimated by proxy – as is common in cost-of-illness studies – using the results from a study of the caregiver burden attributable to DS (Campbell et al., 2018; García-Pérez et al., 2021). The overlap in the AS and DS phenotypes has been described elsewhere (Willemsen et al., 2012). Accordingly, results have been presented for the base-case scenario, accompanied by uncertainty ranges and sensitivity analyses. Regardless, the conclusion is unchanged: that the per-parent burden of AS is large. Further research on the absenteeism and presenteeism attributable to AS involving primary data collection from AS parents is needed. Secondly, the analysis only considered the parental productivity impact of AS over a 10-year period. This short time horizon was adopted to moderate the impact of the uncertainty of key data inputs. Another limitation arose from the assumption that parents who worked remained in the same jobs, and that among those who worked, their hourly contributions to GDP were the same as the average for all working Australians. This approach also assumed that the impact of other mechanisms by which parents contributed to the economy, such as through greater purchasing of healthcare goods and services, were negligible. Previous studies have found that the parents of children with AS have had to change careers in order to accommodate their caregiving responsibilities, while others reported that their progression in their chosen career track was stunted (Arora et al., 2020). Furthermore, limitations in the granularity of data available to support the estimation of the parental productivity index meant that the parental productivity impact associated with the ‘lengthy diagnostic odyssey’ commonly experienced by the families of persons with AS was not to be estimated (Wheeler et al., 2017). However, it is anticipated that diagnostic approaches which support reducing the time to a diagnosis of AS could have a flow-on benefit of reducing a component of the parental stress associated with having a child with AS (Baker et al., 2021; Doble et al., 2020; Godler et al., 2022).

In summary, this is the first known study to estimate the total impact of caring for a child with AS on parental productivity, and the PALYs lost by a parental or caregiver population. This lost parental productivity should be considered when determining the supports that should be provided to persons with AS and their families. Improving parents’ ability to participate in the workforce will likely generate significant flow-on benefits to parents’ mental health, financial status, and relationships with their spouse/partner; other children; extended family; and social network, as well as to the broader economy.

## Data Availability

The datasets generated during and/or analysed during the current study are available from the corresponding author on reasonable request.

## Declarations

### Ethics approval

The study was approved by the Royal Children’s Hospital Human Research Ethics Committee (reference number 33066) and the Monash University Human Research Ethics Committee (reference number 2021-25930-55113). All parents/caregivers provided written informed consent and participants who were deemed cognitively able also provided written informed consent. The authors certify that the study was performed in accordance with the ethical standards as laid down in the 1964 Declaration of Helsinki and its later amendments or comparable ethical standards.

### Consent for publication

Not applicable.

## Acknowledgements

The authors would like to thank all the study participants and their families for their contributions to the study.

## Author contributions

Conceptualization: S.L.H., Data collection: E.K.B., Formal Analysis: S.L.H., Investigation: S.L.H., Methodology: S.L.H., Project Administration: S.L.H., Supervision: D.L., Writing – Original Draft: S.L.H., Writing – Review & Editing: D.L., D.E.G., E.K.B., S.L.H.

## Abbreviations

AS: Angelman syndrome
CHEERS: Consolidated Health Economic Evaluation Reporting Standards
CSRI: Client Service Receipt Inventory
DS: Dravet syndrome
FTE: full-time equivalent
PALY: productivity-adjusted life year
PDMA: Participant Developmental and Medical Anamnesis
WPAI: Work Productivity and Activity Impairment

