## Appendices for "Estimating the impact of Angelman syndrome on parental productivity in Australia using productivity-adjusted life years"

### Table A.1 Key cost-of-illness model inputs

| Parameter | Value (units) |
|---|---|
| **AS prevalence** | |
| AS population prevalence, lower scenario | 0.0012 (percentage of the population, %) (Jørgensen et al., 2019) |
| AS population prevalence, base-case scenario | 0.0025 (percentage of the population, %) (Thomson et al., 2006) |
| AS population prevalence, upper scenario | 0.012 (percentage of the population, %) (Godler et al., 2022) |
| Disabled population age and sex distribution | Age and sex dependent (*Disability, Ageing and Carers*, 2018) |
| AS population life expectancy | 70.00 (years of age) (Coppus, 2013; Dagli et al., 2011) |
| **Parental prevalence** | |
| Average parental age at child's birth | Age and sex dependent (*Births*, 2019) |
| **PALYs lost** | |
| Average paternal workforce participation | 97.27 (percentage of fathers, %) (Baker et al., 2021) |
| Average maternal workforce participation | 47.42 (percentage of mothers, %) (Baker et al., 2021) |
| Attributable absenteeism | 7.15 (hours per week) (Campbell et al., 2018) |
| Attributable presenteeism | 58.00 (percentage of total working time, %) (Campbell et al., 2018) |
| Average workforce participation rate | Age and sex dependent (percentage of the population, %) (*Characteristics of Employment*, 2020) |

| Parameter | Value (units) |
|---|---|
| **Cost to society** | |
| Gross domestic product (GDP) per hour worked per person | 100.00 (Australian dollars, AUD$) (*National Income, Expenditure and Product*, 2020) |
| Proportion full-time equivalent (FTE) | Age and sex dependent (percentage of the population, %) (*Characteristics of Employment*, 2020) |
| **Background** | |
| General population, total | 25,365,745.00 (number of people) (*National, State and Territory Population*, 2020) |
| General population, mortality rate | Age and sex dependent (percentage of the population, %) (*Deaths*, 2019) |
| Annual discount rate | 5.00 (percentage per annum, %) (Larg & Moss, 2011) |

**Table A.2 Assumptions underlying the cost-of-illness modelling approach**

| ID | Assumption |
|---|---|
| A1 | The prevalence of AS approximates 1 in 40,000 (base-case scenario), and falls within the range of 1 in 86,250 (lower scenario) to 1 in 8,290 (upper scenario) (Godler et al., 2022; Jørgensen et al., 2019; Thomson et al., 2006) |
| A2 | The age and sex distribution of AS prevalence followed the same distribution as that of the disabled population and the prevalence of AS within each age group was equally distributed |
| A3 | The average life expectancy of persons with AS was 70 years (Coppus, 2013; Dagli et al., 2011) |
| A4 | Each person with AS had one biological mother and one biological father who experienced productivity impacts attributable to AS |
| A5 | The parents of persons with AS had the same average age as all Australians when their child was born |
| A6 | The impact of average maternal and paternal ages not stated were negligible, and ages falling within the 0 to 15 years range could be taken as 15 years of age, and ages falling within the 49 (maternal) or 59 (paternal) years and over range could be taken as 49 or 59 years of age, respectively |
| A7 | The compound annual growth rate observed in average maternal and paternal age could be applied retrospectively to estimate the average maternal and paternal age for years where data is not available (i.e. from 1919 to 1974) |
| A8 | The mortality rate of persons with AS had a negligible impact on the AS attributable productivity impacts experienced by parents |
| A9 | The mortality rate of the parents of persons with AS was the same as that of the general population, meaning there was no parental mortality attributable to having a child with AS; therefore, the parental and control life years lived were the same |
| A10 | The average workforce participation reported by the small sample size of parents of persons who participated in the natural history study of chromosome 15 imprinting disorders (Baker et al., 2018) was reflective of the average workforce participation of the parents of persons with AS across all age groups included in the analysis |
| A11 | The impact of the following data cleaning decisions on the parental productivity index estimated was negligible: blank responses were not included, siblings were not removed, data reported for partners was taken as the other biological parent, parental age was estimated by summing the age of the child at collection with the time since collection to the mid-point of the initial cycle year (i.e. 1 July 2019) |

| ID | Assumption |
|---|---|
| A12 | The parents of persons with AS who worked part-time had the same average FTE rate as the part-time general population |
| A13 | There were 52 paid weeks in a year, 48 working weeks in a year, 5 working days in a week and 7.5 working hours in a day |
| A14 | The level of parental absenteeism and presenteeism attributable to Dravet syndrome was a reasonable approximation of the parental absenteeism and presenteeism attributable to AS |
| A15 | The productivity index estimated for the parents of persons with AS was a reasonable approximation of the average productivity index across all age groups included in the analysis |
| A16 | The general population employment data approximates the average workforce participation of the control population |
| A17 | Employment statistics for the age group 65 and over were mostly attributable to persons falling within the 65 to 69 age group |
| A18 | The hourly contributions to GDP made by parents who worked were the same as the average for all working Australians |
| A19 | The impact of other mechanisms by which parents contributed to the economy, such as through greater purchasing of healthcare goods and services, were negligible |
| A20 | Productivity-adjusted life years (PALYs) and GDP contributions could be discounted at 5% per annum |

### *References*

Baker, E. K., Arora, S., Amor, D. J., Date, P., Cross, M., O'Brien, J., Simons, C., Rogers, C., Goodall, S., Slee, J., Cahir, C., & Godler, D. E. (2021). The Cost of Raising Individuals with Fragile X or Chromosome 15 Imprinting Disorders in Australia. *Journal of Autism and Developmental Disorders*, *0123456789*. https://doi.org/10.1007/s10803-021-05193-4

Baker, E. K., Godler, D. E., Bui, M., Hickerton, C., Rogers, C., Field, M., Amor, D. J., & Bretherton, L. (2018). Exploring autism symptoms in an Australian cohort of patients with Prader-Willi and Angelman syndromes. *Journal of Neurodevelopmental Disorders*, *10*(1), 24. https://doi.org/10.1186/s11689-018-9242-0

*Births*. (2019). Australian Bureau of Statistics. https://www.abs.gov.au/statistics/people/population/births-australia/latest-release#data-download

Campbell, J. D., Whittington, M. D., Kim, C. H., VanderVeen, G. R., Knupp, K. G., & Gammaitoni, A. (2018). Assessing the impact of caring for a child with Dravet syndrome: Results of a caregiver survey. *Epilepsy & Behavior*, *80*, 152–156. https://doi.org/10.1016/j.yebeh.2018.01.003

*Characteristics of Employment*. (2020). Australian Bureau of Statistics. https://www.abs.gov.au/statistics/labour/earnings-and-work-hours/characteristics-employment-australia/latest-release

Coppus, A. M. W. (2013). People with intellectual disability: What do we know about adulthood and life expectancy? *Developmental Disabilities Research Reviews*, *18*(1), 6–16. https://doi.org/10.1002/ddrr.1123

Dagli, A., Buiting, K., & Williams, C. A. (2011). Molecular and Clinical Aspects of Angelman Syndrome. *Molecular Syndromology*, *2*(3–5), 100–112. https://doi.org/10.1159/000328837

*Deaths*. (2019). Australian Bureau of Statistics. https://www.abs.gov.au/statistics/people/population/deaths-australia/latest-release

*Disability, Ageing and Carers*. (2018). Australian Bureau of Statistics. https://www.abs.gov.au/statistics/health/disability/disability-ageing-and-carers-australia-summary-findings/latest-release#data-download

Godler, D. E., Ling, L., Gamage, D., Baker, E. K., Bui, M., Field, M. J., Rogers, C., Butler, M. G., Murgia, A., Leonardi, E., Polli, R., Schwartz, C. E., Skinner, C. D., Alliende, A. M., Santa Maria, L., Pitt, J., Greaves, R., Francis, D., Oertel, R., … Amor, D. J. (2022). Feasibility of Screening for Chromosome 15 Imprinting Disorders in 16579 Newborns by Using a Novel Genomic Workflow. *JAMA Network Open*, *5*(1), 1–13. https://doi.org/10.1001/jamanetworkopen.2021.41911

Jørgensen, I. F., Russo, F., Jensen, A. B., Westergaard, D., Lademann, M., Hu, J. X., Brunak, S., & Belling, K. (2019). Comorbidity landscape of the Danish patient population affected by chromosome abnormalities. *Genetics in Medicine*, *21*(11), 2485–2495. https://doi.org/10.1038/s41436-019-0519-9

Larg, A., & Moss, J. R. (2011). Cost-of-Illness Studies. *PharmacoEconomics*, *29*(8), 653–671. https://doi.org/10.2165/11588380-000000000-00000

*National, state and territory population*. (2020). Australian Bureau of Statistics. https://www.abs.gov.au/statistics/people/population/national-state-and-territory-population/jun-2020#data-download

*National income, expenditure and product*. (2020). Australian Bureau of Statistics. https://www.abs.gov.au/statistics/economy/national-accounts/australian-national-accounts-national-income-expenditure-and-product/latest-release#data-download

Thomson, A., Glasson, E., & Bittles, A. (2006). A long-term population-based clinical and morbidity profile of Angelman syndrome in Western Australia: 1953–2003. *Disability and Rehabilitation*, *28*(5), 299–305. https://doi.org/10.1080/09638280500190631
